# Performance of qualitative and quantitative antigen tests for SARS-CoV-2 in early symptomatic patients using saliva

**DOI:** 10.1101/2020.11.06.20227363

**Authors:** Isao Yokota, Takayo Sakurazawa, Junichi Sugita, Sumio Iwasaki, Keiko Yasuda, Naoki Yamashita, Shinichi Fujisawa, Mutsumi Nishida, Satoshi Konno, Takanori Teshima

## Abstract

**Background:** The rapid detection of severe acute respiratory syndrome coronavirus 2 (SARS-CoV-2) is an urgent need for the prevention and containment of disease outbreaks in communities. Although the gold standard is polymerase chain reaction (PCR), antigen tests such as immunochromatographic assay (ICA) and chemiluminescent enzyme immunoassay (CLEIA) that can yield results within 30 minutes.

**Methods:** We evaluated performance of ICA and CLEIA using 34 frozen PCR-positive specimens (17 saliva and 17 nasopharyngeal swab) and 307 PCR-negative samples.

**Results:** ICA detected SARS-CoV-2 in only 14 (41%) samples, with positivity of 24% in saliva and 59% in NPS. Notably, ICA detected SARS-CoV-2 in 5 (83%) of 6 samples collected within 4 days after symptom onset. CLEIA detected SARS-CoV-2 in 31 (91%) samples, with positivity of 82% in saliva and 100% in NPS. CLEIA was negative in 3 samples with low viral load by PCR.

**Conclusions:** These results suggest that use of ICA should be limited to earlier time after symptom onset and CLEIA is more sensitive and can be used in situations where quick results are required.

## Introduction

Rapid detection of the novel coronavirus severe acute respiratory syndrome coronavirus 2 (SARS-CoV-2) is critical for the prevention and containment of coronavirus disease-19 (COVID-19) outbreaks in communities and hospitals. Currently, the “gold standard” of viral detection is quantitative reverse transcriptase polymerase chain reaction (PCR) using nasopharyngeal swab samples (NPS) [1, 2]. NPS sampling requires specialized medical personnel with protective equipment, posing risks of viral transmission to healthcare workers, and false-negative results may occur due to deficiency in sampling technique[2, 3]. Self-collected saliva can be as effective as traditional nasal swabbing, making a major step for a type of screening, which is much faster, and less instructive and expensive[4-6].

Although PCR is highly accurate and reliable, it is time-consuming as a screening test. Viral antigen detection tests such as a rapid antigen test (immunochromatographic assay, ICA) and chemiluminescent enzyme immunoassay (CLEIA) are much more easy and yield results quickly than PCR[7-11]. In this study, we evaluated the utility of ICA and CLEIA in comparison with PCR using self-collected saliva and NPS.

## Methods

### Samples

We screened 34 SARS-CoV-2 positive samples (17 NPS and 17 saliva samples) as established by PCR and 307 negative saliva samples. All positive samples were obtained from symptomatic patients with COVID-19 and all the negative samples were obtained from asymptomatic persons at screening. All the samples had been frozen and thawed before analysis. This study was approved by the Institutional Ethics Board (Hokkaido University Hospital Division of Clinical Research Administration Number: 020-0116), and informed consent was obtained from all patients.

Saliva and NPS samples were collected as previously described[12]. Saliva samples were diluted 4-fold with phosphate buffered saline and centrifuged at 20,000 × g for 5 min at 4°C to remove cells and debris[12]. Following thawing, samples were centrifuged at 2,000 × g for 5 min at 4°C to remove cells and debris. Each sample was divided for application to four testing.

### PCR

RNA was extracted using QIAamp Viral RNA Mini Kit (QIAGEN, Hilden, Germany). PCR tests were performed as described[12], according to the manual by National Institute of Infectious Diseases (NIID, https://www.niid.go.jp/niid/images/epi/corona/2019-nCoVmanual20200217-en.pdf). Briefly, 5uL of the extracted RNA was used as a template. One step PCR was performed using One-Step Real-Time RT-PCR Master Mixes (Thermo Fisher Scientific, Waltham, USA) and Real Time PCR System (Thermo Fisher Scientific). The cycle threshold (Ct)-values were obtained using N2 primers (NIID_2019-nCOV_N_F2, NIID_2019-nCOV_N_R2) and a probe (NIID_2019-nCOV_N_P2).

### Antigen tests

ICA was performed using Espline SARS-CoV-2 (Fujirebio, Tokyo, Japan) according to the manufacturer’s instructions. In brief, samples were mixed with the sample preparation mixture. The mixture (200 μl) and 2 drops of buffer was added to the ample port and the results were interpreted after incubation for 30 minutes.

Lumipulse SARS-CoV-2 Ag kit® (Fujirebio, Tokyo, Japan) is a sandwich CLEIA using monoclonal antibodies that recognize SARS-CoV-2 N-Ag on LUMIPULSE G1200 automated machine (Fujirebio). 100 µL of saliva diluted with PBS was analyzed to measure N-Ag levels according to the manufacturer’s instructions. In brief, the treatment solution and the specimen were consecutively aspirated using a single-use tip and was dispensed into suspension of magnetic beads coated with the monoclonal antibody to SARS-CoV-2 N-Ag. After 10-minute incubation followed by a wash-step, alkaline phosphatase-conjugated anti-SARS-CoV-2 monoclonal antibody were added and incubated for another 10 minutes. After a second wash-step, AMPPD substrate solution was added and developed for 5 minutes. The amounts of SARS-CoV-2 N-Ag were calibrated with the developed chemiluminescence signals. The calibrator of this assay contains the recombinant SARS-CoV-2 N antigen standardized with the recombinant SARS-CoV-2 antigen. Antigen levels of 0.67 pg/mL or greater were defined as positive.

### Statistical analysis

Using positive samples detected by PCR, the proportion of positivity in each test was calculated with 95% confidential interval (CI). The values detected by CLEIA and ICA test were plotted against Ct value of PCR test. Kendall’s coefficient of concordance W was calculated as nonparametric intraclass correlation coefficient. The values were also plotted against the days from symptom onset in order to determine the trend over time. Histograms of antigen concentration in the CLEIA test are drawn for both positive and negative samples. All statistical analyses were conducted by R 4.0.2 and Clopper-Pearson exact confidence interval was used for a proportion.

## Results

SARS-CoV-2 positive samples included 17 NPS and 17 self-collected saliva. Median time of sampling was 9 days (range, 2-14 days) after symptom onset. In all 34 virus positive samples (17 saliva and 17 NPS) as established by PCR before freezing, PCR positivity was again confirmed after thawing. ICA detected viral antigens in only 14 (41%, 95%CI: 25–59%) samples (Table 1). Particularly, the virus was positive in only 24% (95%CI: 7–50%) in saliva in contrast to 59% (95%CI: 33–82%) positivity in NPS. It should be noted that ICA was positive in 5 (83%, 95%CI: 36-100%) out of 6 samples collected within 4 days after symptom onset and in 9 (32%, 95%CI: 16-52%) of 28 samples collected thereafter. In NPS, it was positive in 9 (82%, 95%CI: 48-98%) out of 11 samples collected within 10 days after symptom onset, but in 1 (17%, 95%CI: 0-64%) of 6 samples collected thereafter. In saliva, all 3 samples collected at 2-4 days after symptom onset were positive, but only 1 (7%, 95%CI: 0-34%) of 14 samples collected thereafter were positive. Raw data are shown in Supplement 1.

**Table 1.** Diagnostic results in positive specimens diagnosed by RT-PCR.

| Test | Positive (% , 95% confidence interval) |  |  |
| --- | --- | --- | --- |
|  | Total<br>(n=34) | Saliva<br>(n=17) | NPS<br>(n=17) |
| ICA | 14<br>(41% , 25–59%) | 4<br>(24% , 7–50%) | 10<br>(59% , 33–82%) |
| CLEIA | 31<br>(91% , 76–98%) | 14<br>(82% , 57–96%) | 17<br>(100% , 80–100%) |

On the other hand, CLEIA yielded 91% (95%CI: 76–98%) positivity, with 82% (95%CI: 57–96%) positivity in saliva and 100% (95%CI: 80–100%) positivity in NPS. However, 3 out of 34 samples were CLEIA-negative. These samples were all saliva collected at 7, 12, and 14 days after symptom onset, and Ct values were 32.4-33.8 by PCR.

A scatter plot of antigen concentrations with CLEIA against Ct values of PCR test is shown in Figure 1a. Kendall’s coefficient of concordance was 0.91, indicating high correlation between CLEIA and PCR in both saliva and NPS. ICA positivity tended to have higher viral loads of PCR (Ct values: 21.6 [interquartile range, IQR]: 19.1–23.3) in ICA positive vs. 29.6 [IQR: 28.0–30.9] in ICA negative), but many PCR positive samples were ICA negative, particularly in saliva (Figure 1b). Antigen concentrations determined by CLEIA declined over time after symptom onset (Figure 2a). Similarly, frequency of ICA positivity decreased over time (Figure 2b).

**Figure 1.**
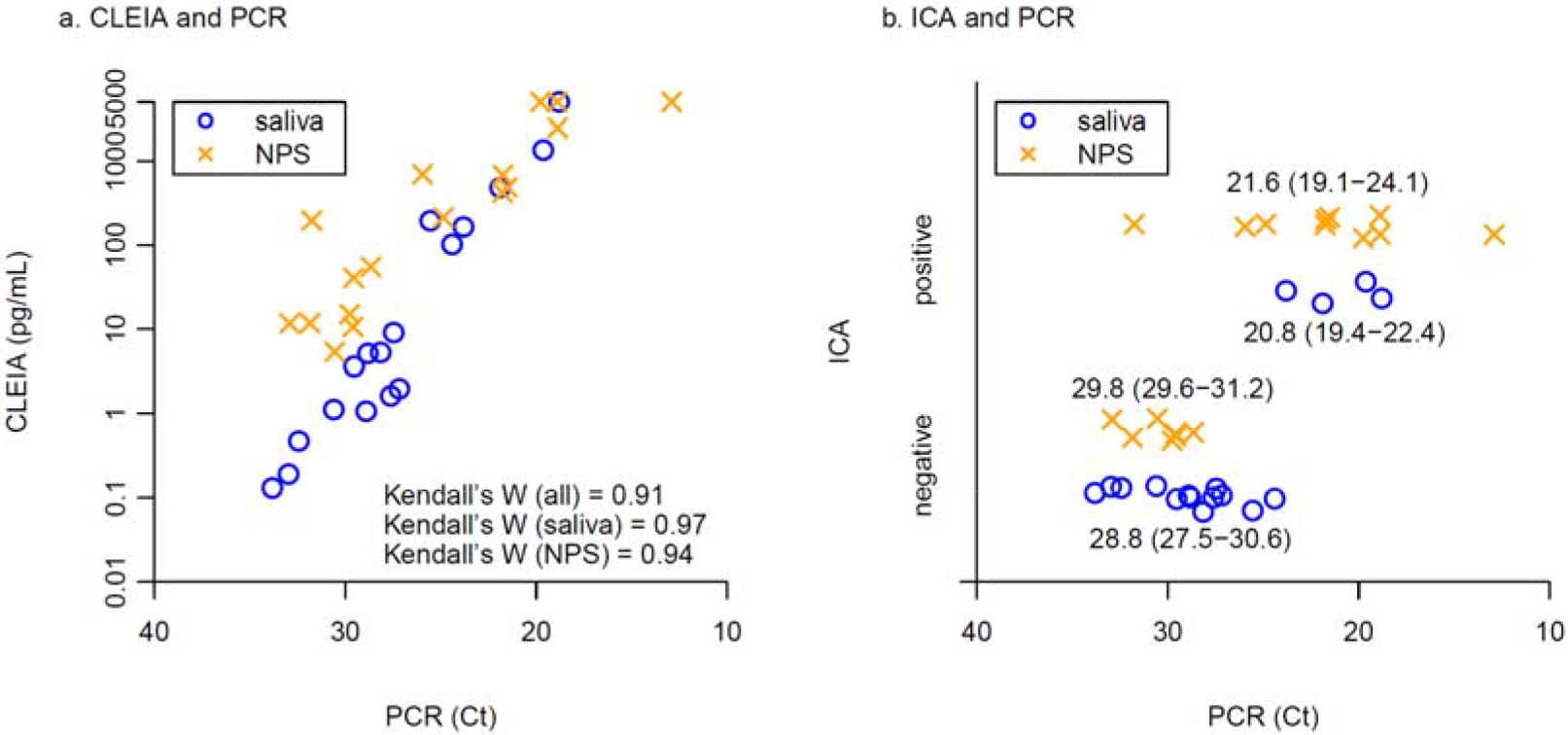
Viral loads by CLEIA and ICA in comparison with PCR. A scatter plot shows relationships between (a) antigen concentrations of CLEIA and Ct values of PCR, and (b) ICA positivity/negativity and Ct values of PCR. Blue circles and yellow crosses represent saliva and NPS samples, respectively. Median and interquartile range of Ct values was shown in (b).

**Figure 2.**
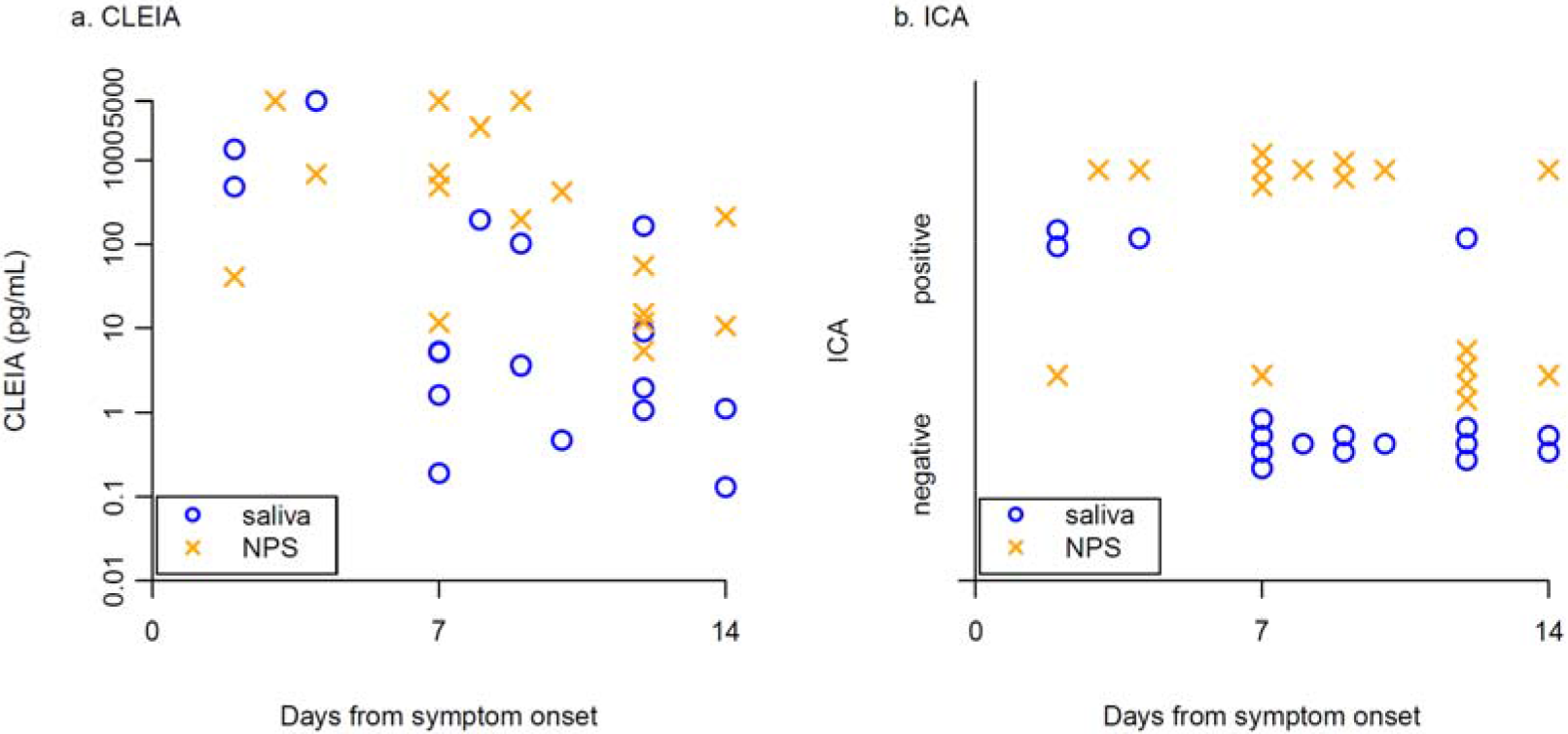
Viral loads by CLEIA and ICA according to time from symptom onset. A scatter plot shows relationship between days from symptom onset and (a) antigen concentrations of CLEIA, (b) Ct values of PCR. Blue circles and yellow crosses represent saliva and NPS samples, respectively.

The distribution of antigen concentrations determined by CLEIA in 34 PCR-positive and 307 PCR-negative samples is shown in Figure 3. The median (IQR) antigen concentration was 48.2 (5.2 – 486.7) pg/mL in PCR-positive specimens and 0.03 (0.01 – 0.09) pg/mL in PCR-negative specimens. The maximum of antigen concentration in PCR-negative specimens was 24.23 pg/mL. Raw data are shown in Supplement 2.

**Figure 3.**
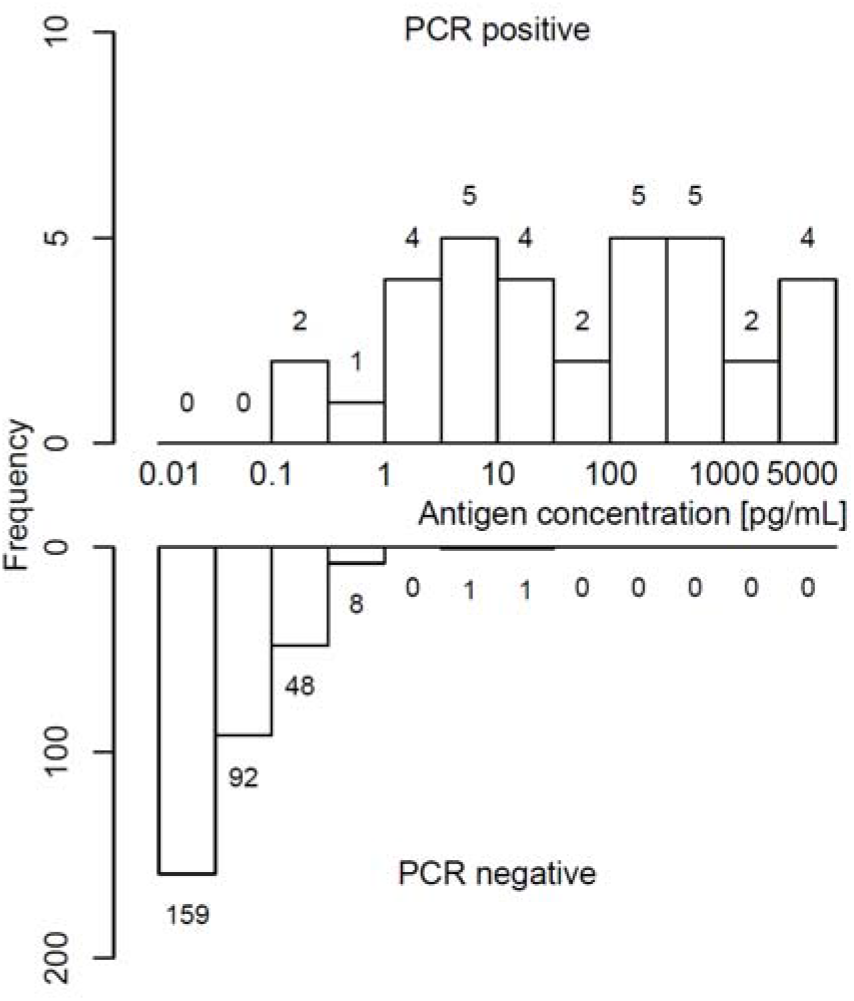
Histogram of antigen concentration by the diagnostics of the PCR test. Upper and lower panels show frequency of PCR-positivity and PCR-negativity, respectively, according to CLEIA antigen concentrations. Frequencies in each range were shown above/below histogram.

## Discussion

Our results suggest that ICA could be used only within 4 days after symptom onset using both NPS and self-collected saliva. However, ICA is not reliable in samples collected thereafter with significant concerns of high false-negative rates. Sensitivity of ICA markedly decreased over time after symptom onset, particularly in saliva. It is well documented that SARS-CoV-2 tends to persist longer in NPS than in saliva[12, 13]. It is thus reasonable to speculate that lower sensitivity of saliva ICA is due to late sampling rather than the difference in antigen load between saliva and NPS. Overall, sensitivity of ICA using NPS was 59%, which was similar to 51-67.5% sensitivity of rapid antigen test using NPS for influenza[14-17]. It should be noted that antigen testing of influenza using NPS is also recommended to perform within 3 days after symptom onset[17]. ICA is suitable for point-of-care testing and therefore these results should be confirmed in larger cohort.

In contrast, CLEIA is much more reliable and accurate in both NPS and saliva than ICA, with high correlation observed between antigen concentrations and RNA load by PCR. However, there were three PCR-positive but CLEIA-negative samples. These samples were all saliva collected at 7, 12, and 14 days after symptom onset. We therefore recommend using saliva for CLEIA only in patients who developed symptoms within a week. Moreover, in these samples, Ct values were 32.4-33.8 by PCR. A “positive” PCR result does not necessarily indicate presence of live virus[18], and recent studies showing patients with Ct values above 33-34 by PCR not to be contagious[19, 20]. Vice versa, there were 2 (0.65%) of 307 samples were PCR-negative but CLEIA-positive with high antigen concentrations of 8.45 and 24.23 pg/mL. Reexamination of these specimens confirmed CLEIA-positivity. This could reflect a false positive CLEIA, but the possibility of false negative PCR result cannot be completely ruled out and the clinical implication of this discrepancy remains to be elucidated.

Measuring antigen concentrations by CLEIA is a novel approach to viral detection with equivalent utility compared to PCR. To improve diagnostic accuracy, two-step testing with initial CLEIA and secondary nucleic acid amplification tests (NAAT) is reasonable. Thus, the great advantage of having a rapid and accurate CLEIA is the ability for primary screening to establish a “gray zone” reserving definitive diagnosis by confirmatory NAAT testing. Reverse transcriptase loop-mediated isothermal amplification (LAMP)[7] has become the second most common NAAT after PCR with several advantages over PCR: rapid turn-around time within 30 minutes, ease of implementation, and potential utility at point of care using a simple device[6, 9, 21-25]. We recently reported that LAMP had equivalent efficacy to PCR using saliva in both asymptomatic persons and symptomatic patients [6]. Thus, LAMP is currently being used at the international airport quarantine as the confirmatory NAAT testing of CLEIA in Japan.

## Conclusions

Antigen detection of SARS-CoV-2 yield results quickly. However, use of a rapid antigen test should be limited to within a few days after symptom onset. A quantitative antigen test using both nasopharyngeal and saliva specimens shows high concordance with PCR and can be used in situations where quick results are required.

## Data Availability

All data are included in Supplements.

## List of abbreviations

SARS-CoV-2: severe acute respiratory syndrome coronavirus 2
PCR: polymerase chain reaction
LAMP: loop-medicated isothermal amplification
ICA: immunochromatographic assay
CLEIA: chemiluminescent enzyme immunoassay
COVID-19: coronavirus disease-19
NPS: nasopharyngeal swab samples
NAAT: nucleic acid amplification test
NIID: National Institute of Infectious Diseases
CI: confidential interval
Tp: time for detecting positive results
IQR: interquartile range

## Declarations

### Ethical approval and consent to participate

This study was approved by the Institutional Ethics Board (Hokkaido University Hospital Division of Clinical Research Administration Number: 020-0116), and informed consent was obtained from all patients. This study was adhered to relevant guidelines and regulations.

### Consent for publication

Not applicable

### Availability of data and materials

The datasets used in this study are available from the corresponding author on reasonable request.

### Authors’ contributions

Study design: IY, JS, SF, MN, TT; Data collection: SK; Data analysis: TS, SI, KY, NY, SF; Writing: IY, TS, TT

