## Supplemental Table for "Performance of qualitative and quantitative antigen tests for SARS-CoV-2 in early symptomatic patients using saliva"

Supplement 1. Raw data in PCR-positive specimens

| Days after symptom onset | specimen | Ct value | Tp value (sec) | CLEIA (pg/mL) | ICA |
|---|---|---|---|---|---|
| 2 | NPS | 29.56192 | 586 | 40.87 | negative |
| 3 | NPS | 12.90119 | 301 | 5000 | positive |
| 4 | NPS | 21.74256 | 391 | 685.64 | positive |
| 7 | NPS | 18.89330 | 346 | 5000 | positive |
| 7 | NPS | 21.52387 | 391 | 487.16 | positive |
| 7 | NPS | 25.95481 | 451 | 699.01 | positive |
| 7 | NPS | 32.91834 | 661 | 11.62 | negative |
| 8 | NPS | 18.88139 | 436 | 2484.37 | positive |
| 9 | NPS | 19.76842 | 346 | 5000 | positive |
| 9 | NPS | 31.75552 | 646 | 196.65 | positive |
| 10 | NPS | 21.75409 | 391 | 422.62 | positive |
| 12 | NPS | 31.85005 | 526 | 11.77 | negative |
| 12 | NPS | 30.54592 | 601 | 5.34 | negative |
| 12 | NPS | 29.77066 | 511 | 15.02 | negative |
| 12 | NPS | 28.65936 | 556 | 55.43 | negative |
| 14 | NPS | 24.86924 | 406 | 213.09 | positive |
| 14 | NPS | 29.59635 | 556 | 10.62 | negative |
| 2 | saliva | 19.62641 | 391 | 1357.72 | positive |
| 2 | saliva | 21.89011 | 376 | 485.26 | positive |
| 4 | saliva | 18.79502 | 511 | 5000 | positive |
| 7 | saliva | 28.14606 | 526 | 5.26 | negative |
| 7 | saliva | 32.96928 | 751 | 0.19 | negative |
| 7 | saliva | 27.62087 | 541 | 1.63 | negative |
| 7 | saliva | 28.81459 | 571 | 5.12 | negative |
| 8 | saliva | 25.55043 | 436 | 194.68 | negative |
| 9 | saliva | 24.40270 | 481 | 102.21 | negative |
| 9 | saliva | 29.54057 | 526 | 3.62 | negative |
| 10 | saliva | 32.43248 | 496 | 0.47 | negative |
| 12 | saliva | 28.90566 | 571 | 1.07 | negative |
| 12 | saliva | 27.46050 | 586 | 9.06 | negative |
| 12 | saliva | 27.17160 | 616 | 1.98 | negative |
| 12 | saliva | 23.80877 | 496 | 164.85 | positive |
| 14 | saliva | 33.81876 | 601 | 0.13 | negative |
| 14 | saliva | 30.60193 | 601 | 1.12 | negative |

Supplement 2. Antigen concentration by CLEIA in PCR-negative specimens

| Antigen concentration (pg/mL) | Frequency | Antigen concentration (pg/mL) | Frequency |
|---|---|---|---|
| 0.01 | 122 | 0.19 | 1 |
| 0.02 | 18 | 0.20 | 2 |
| 0.03 | 19 | 0.21 | 2 |
| 0.04 | 16 | 0.22 | 1 |
| 0.05 | 14 | 0.23 | 2 |
| 0.06 | 12 | 0.24 | 1 |
| 0.07 | 6 | 0.25 | 1 |
| 0.08 | 19 | 0.27 | 1 |
| 0.09 | 11 | 0.28 | 1 |
| 0.10 | 14 | 0.30 | 1 |
| 0.11 | 5 | 0.34 | 2 |
| 0.12 | 3 | 0.37 | 1 |
| 0.13 | 6 | 0.39 | 1 |
| 0.14 | 7 | 0.40 | 2 |
| 0.15 | 3 | 0.60 | 1 |
| 0.16 | 6 | 0.63 | 1 |
| 0.17 | 4 | 8.45 (4.78)* | 1 |
| 0.18 | 1 | 24.23 (16.07)* | 1 |

* The antigen concentration at re-test was shown in parenthesis.
